# Experiences of living with mental health problems during the COVID-19 pandemic in the UK: a coproduced, participatory qualitative interview study

**DOI:** 10.1101/2020.11.03.20225169

**Authors:** Steven Gillard, Ceri Dare, Jackie Hardy, Patrick Nyikavaranda, Rachel Rowan Olive, Prisha Shah, Mary Birken, Una Foye, Josephine Ocloo, Ellie Pearce, Theodora Stefanidou, Alexandra Pitman, Alan Simpson, Sonia Johnson, Brynmor Lloyd-Evans

## Abstract

**Purpose:** Research is beginning to quantify the impact of COVID-19 on people with pre-existing mental health conditions. Our paper addresses a lack of in-depth qualitative research exploring their experiences and perceptions of how life has changed at this time.

**Methods:** We used qualitative interviews (N=49) to explore experiences of the pandemic for people with pre-existing mental health conditions. In a participatory, coproduced approach, researchers with lived experiences of mental health conditions conducted interviews and analysed data as part of a multi-disciplinary research team.

**Results:** Existing mental health difficulties were exacerbated for many people. People experienced specific psychological impacts of the pandemic, struggles with social connectedness, and inadequate access to mental health services, while some found new ways to cope and connect to community. New remote ways to access mental health care, including digital solutions, provided continuity of care for some but presented substantial barriers for others. People from black and ethnic minority (BAME) communities experienced heightened anxiety, stigma and racism associated with the pandemic, further impacting their mental health.

**Conclusion:** There is a need for evidence-based solutions to achieve accessible and effective mental health care in response to the pandemic, especially remote approaches to care. Further research should explore the long-term impacts of COVID-19 on people with pre-existing mental health conditions. Particular attention should be paid to understanding inequalities of impact on mental health, especially for people from BAME communities.

## Introduction

The negative impacts of the novel coronavirus infectious disease 2019 (COVID-19) pandemic on society include illness, death and bereavement, social isolation, unemployment, disruption to education and loss of access to services and resources resulting from infection control restrictions (‘lockdown’). Like previous pandemics, [1] COVID-19 has caused psychological distress for many, [2] with steep rises in mental distress among young people and women in particular, [3] and increased incidence of psychosis having been reported. [4]

The effects of the pandemic may exacerbate existing inequalities. People from Black, Asian and Minority Ethnic (BAME) communities have been found to have higher rates of hospitalization and mortality from the virus in the UK and USA, [5,6] with higher levels of socio-economic disadvantage in BAME communities potentially contributing to an increased risk of infection. [7] Impacts on mental health may also be unequal. Living in poor housing during extended spells of ‘lockdown’ has been associated with greater levels of depression. [8] In a UK survey, BAME respondents were most likely to report that financial worries and problems with housing and employment were having a negative impact on their mental health as a result of the pandemic. [9]

An evidence synthesis from the earliest phase of the pandemic [10] suggested that many people with existing mental health conditions experienced increased difficulties regarding loneliness and isolation, loss of routine, conflicts and abuse within the family including domestic violence, lack of access to services, challenges with transition to remote care, and infection control in inpatient environments. People with existing mental health conditions may be more adversely affected by the pandemic than the general population, with higher rates of COVID-19 infection and concerns that factors such as comorbidity and substance use may increase susceptibility to a severe illness course. [11] An Australian survey found that people with self-reported mood disorder experienced higher levels of psychological distress and adverse changes to lifestyle as a result of the pandemic than individuals reporting no mental disorder. [12]

Mental health services face increased demand as a result of the pandemic [13] in the challenging context of reductions in capacity and the requirements of infection control and safe service provision. [14,15] A survey of 2,180 mental health staff in the UK [16] found infection control and lack of meaningful activity for service users were major concerns for inpatient and residential settings, while staff in community services typically struggled with rapid adaptation to new ways of working, including remote care, and lack of other community and voluntary sector services to refer into. Increased workload, introducing telehealth technologies and altered patient/provider interactions have been identified as key challenges by US mental health staff. [17] Individual approaches to self-management and increased use of peer or community support by people with mental health conditions have been reported. [10] However, a lived experience commentary on this paper questioned the sustainability of such self-management.

To help understand and address these challenges, rapid and collaborative mental health research has been called for. [18] People living with mental health difficulties have been active in writing personal accounts of the pandemic. [19] However, very little work has systematically investigated the views and experiences of the pandemic of people already living with mental health conditions and receiving mental health care. This perspective is crucial to inform the immediate and ongoing mental health service response to COVID-19. Our study addresses these needs for knowledge by exploring, in depth, the experiences of people with pre-existing mental health problems during the COVID-19 pandemic.

## Methods

### Study design

We used a participatory, coproduction approach to conducting a qualitative interview study. [20] Coproduction of qualitative research has been described as enabling shared decision-making, alongside reflection on the interpretive process, across researchers working from experiential, clinical and academic perspectives. [21] The longstanding tradition of community participatory research encourages community-based researchers to actively use their lived experience in shaping the research process, [22] and can attend to health inequalities through embedding cultural context into the research. [23] Methods were adapted to respond to the challenges of working remotely during the pandemic.

Ethical approval for a study focusing on loneliness was obtained from the UCL Research Ethics Committee on 19/12/2019 (ref: 15249/001). An amended topic guide covering experiences of COVID-19 was approved on 04/05/2020. Findings relevant to experiences of the pandemic are reported in this paper.

### Team

The fifteen authors met weekly to manage the project and collectively wrote this paper. This group included six researchers who worked from a lived experience perspective, and 10 (including three clinical academics) who worked in university research staff roles from a range of disciplinary perspectives. Twelve authors were women and three from BAME backgrounds. Eight additional lived experience researchers (LERs), together with five LER authors, contributed to developing topic guides, conducted all interviews and undertook initial coding of interviews. All thirteen LER interviewers had personal experiences of using mental health services and/or mental distress, and worked in university research roles, or were members of research advisory groups or were involved in service user or community organisations. Eleven of the LERs were female and five came from BAME backgrounds.

### Sampling and recruitment strategy

The study population was adults who self-identified as having had experiences of mental health difficulties that preceded the pandemic. We sampled purposively to achieve diversity regarding participants’ diagnoses, use of mental health services, and demography regarding age, gender, ethnicity and sexuality, and from rural and urban areas. We reviewed our sample during recruitment and targeted further recruitment to ensure diversity.

We aimed to recruit 40-50 participants through community organisations, mental health networks and social media, and approached organisations working specifically with BAME communities using targeted recruitment materials and images. Researchers responded to potential participants to check eligibility, provide a participant information sheet, answer questions and set up interviews, where informed consent was given.

### Data collection

Interviews were conducted by LERs using videoconferencing or freephone options within the Microsoft Teams application, with a second study researcher present to support recording and saving the interview in password protected university files. Audio-recorded, verbal informed consent was taken before the interview. We developed and used a semi-structured interview schedule, building on a previous study on loneliness and informed by emerging literature on COVID-19 and mental health, [10,16] and lived experience within the research team. Questions explored experiences of the pandemic, including its impact on people’s mental health and mental health care. The study team developed a guide to online and telephone mental health resources from national and local organisations, which was shared with all participants to provide signposting to support following the interview. Participants were offered a supportive follow-up telephone call or e-mail.

Training for LERs was provided by university researchers through an online workshop. A weekly lived experience reflective space provided LERs with emotional support and space to discuss the research process, peer-facilitated by four experienced LERs.

### Analysis

We sought to integrate experiential knowledge into the interpretive process. Following an online training session delivered by members of the study team, nine LERs each undertook preliminary analysis of an interview they had conducted using general principles of thematic analysis, [24] listening back to recordings and charting a maximum of five emerging thematic ideas per interview onto a coding matrix, capturing interviewees’ experiences of mental health and COVID-19.

Through online workshops, we produced a preliminary coding framework, refining and amalgamating thematic ideas from individual matrices where these were meaningfully similar while retaining idiosyncratic thematic ideas. LERs used the preliminary framework to code all interviews, listening back to interviews and charting quotes and analytical memos. At a further workshop, we finalised the framework, adding and modifying themes as necessary.

Results were produced through an iterative process of analytical writing, [25,26] with university researchers and LERs working together in small groups to write analytical narrative around exemplary quotes, before refining each theme through team discussion and rewriting.

## Results

We recruited 49 people: most (69%) were female, 78% were age 25-54 years, and two-thirds (69%) identified as heterosexual. A majority (53%) were White British, with Black/Black British (14%) and Asian/Asian British (12%) the next most represented ethnic groups. Just over half (53%) lived in London. The majority (86%) reported current or recent mental health service use. See Table 1 for full details.

**Table 1:** Characteristics of participants (N=49)

| Characteristic | Category | Number (%) |
| --- | --- | --- |
| Gender | Female | 34 (69%) |
|  | Male | 15 (31%) |
| Age | 18–24 | 2 (4%) |
|  | 25–34 | 15 (31%) |
|  | 35–44 | 12 (24%) |
|  | 45–54 | 11 (22%) |
|  | 55–69 | 3 (6%) |
|  | ≥70 | 4 (8%) |
|  | Information not available | 2 (4%) |
| Ethnicity | White British | 26 (53%) |
|  | White Other | 5 (10%) |
|  | Mixed/Multiple ethnic groups | 4 (8%) |
|  | Asian/ Asian British | 6 (12%) |
|  | Black / Black British | 7 (14%) |
|  | Other ethnic group | 1 (2%) |
| Sexuality | LGBTQI<sup>1</sup> | 9 (19%) |
|  | Heterosexual | 34 (69%) |
|  | Prefer not to answer or information not available | 6 (12%) |
| Region of UK | North (North East, North West, Yorkshire and Humber) | 9 (18%) |
|  | Midlands (West Midlands, East Midlands) | 6 (12%) |
|  | South (South East, South West, East of England) | 7 (14%) |
|  | London | 26 (53%) |
|  | Wales | 1 (2%) |
| Urban/rural location | City | 38 (76%) |
|  | Town | 9 (18%) |
|  | Village | 2 (4%) |
| Current mental health service use | None, or on waiting list | 12 (24%) |
|  | NHS community mental health services<sup>2</sup> | 23 (47%) |
|  | Inpatient services<sup>3</sup> | 3 (6%) |
|  | GP or primary care counselling | 2 (4%) |
|  | Private sector psychotherapy only | 2 (4%) |
|  | Voluntary sector mental health services only | 4 (8%) |
| Self-reported diagnosis | Personality Disorder | 6 (12%) |
|  | Mood disorders (Depression, Anxiety, PTSD) | 23 (47%) |
|  | Bi-polar Disorder | 5 (10%) |
|  | Schizophrenia/Psychosis | 7 (14%) |
|  | Other (addictions, suicidal thoughts, OCD) | 3 (6%) |
|  | Not stated | 11 (22%) |

1. LGBTQI included: Gay (n=1), Lesbian (n=1), Bisexual (n=6), Pansexual (n=1) 2. Community mental health services included: Community mental health team (n=18), Reablement team (n=1). Therapist (n=2), NHS peer support service (n=2) 3. Inpatient services included: acute inpatient ward (n=2), crisis house (n=1)

Five key themes were identified, each with inter-related subthemes. An analytical summary of each theme is reported here. Additional data can be found in supplementary material.

### Impact of COVID-19 on everyday life and mental health

Lockdown affected participants’ home life, employment and finances. Difficulties included access to essential goods, services, healthcare and leisure activities, and maintaining social distancing. Several people described how their existing mental health difficulties made these challenges more worrying or harder to manage.

*Food shopping, being able to get stuff online was difficult. I don’t drive so [this] was hard. My social anxiety was going up so it affected me going out for food or anything else*. [P32]

Lockdown increased the extent and frequency of contact with other confined household members, which for some explicitly contributed to a deterioration in mental health.

*There’s eight of us in a two-bedroom house which isn’t easy*… *Since lockdown, everything has been contained to within the home. My mental health condition is worsened*. [P17]

Those in work described difficulties getting equipment for remote working or, for one person with physical disabilities, necessary workspace adaptations. Disruption to work or studies reduced people’s independence and caused financial hardships, which affected their mental health.

*The biggest impact [was] almost certainly going bankrupt. Trying to live on universal credit*. [P14]

### Impact of changes to mental healthcare

Some participants described how mental health services seemed slow to adapt and offer support during COVID-19, especially where they thought service provision was already limited. They reported issues with continuity of care, not getting treatment as usual, service changes, and a feeling that professionals were firefighting rather than providing planned care. Some felt they were missing out on care:

*Lockdown has made me feel very angry. I feel the professionals have used it as an excuse to stop offering appointments*. [P4]

*I was seeing her every week and it’s been cut to every 3 weeks [by telephone]* [P25]

Some noted a lack of communication from services, with one worrying that if they were not proactive their care would be discontinued:

*I’m really worried that if I don’t ring them, they will perceive me to be fine and discharge me* … *so I need to ring them and remind them I’m still here and need support*. [P21]

Another participant described how the impact of the pandemic on the psychiatric ward environment became an acute source of distress:

*The ward was really hectic. One day they were discharging everyone, then the next day the consultant came in wearing scrubs … [it] felt a really scary place, I felt unsafe, really wanted to be at home… there was fear on the ward*. [P42]

Community mental health services started to offer remote support by telephone, text and online. While one participant shared a positive experience, others found the level of care deteriorated:

*I had full length appointments with my care co-ordinator over the telephone throughout and they are keeping it really consistent. Every week*. [P44]

*She did text me few times, we keep conversation texting, but it is not good enough really*. [P48]

For some people remote support was challenging, because of their mental health, other conditions, or lack of access to technology:

*For my paranoia, it [video calls] makes it worse, so I tend not to do them*. [P17]

*It’s slightly more tiring, talking online as I’m sensitive to sound, so if there are issues with the sound quality or the words are out of synch or there’s no image, I’m having to focus a lot more because I need visual representation of what’s being said, either subtitling or the image*. [P27]

Privacy could be problematic when attempting to access therapeutic support at home, as not everyone had access to a private space where they could guarantee not being interrupted or overheard:

*I was wary about having my telephone appointment with my psychiatrist over the phone, because of my family situation* … *I don’t talk much about my mental health with them and it was harder talking from home*. [P17]

Some sought help from other sources, including phone calls with their General Practitioner (GP), or turned to the voluntary sector for support that participants felt statutory services were not offering:

*[My] GP has been really helpful, even though often a 5-minute phone call, or 10 minutes, but just having that check in and seeing how I am doing*. [P12]

*I was supposed to receive 1:1 person-centred counselling but I got the appointments with the charity and I would rather have it with them*. [P34]

### Psychological impact

Wide variation in the impact of COVID-19 on individual’s mental health was reported. Many described a decline or large fluctuations resulting from: isolation and loneliness, over-crowded housing; loss of employment, education and social contact; loss of usual coping strategies, structure and purpose; feelings of anger; feeling overwhelmed; confusion around government guidance; sudden changes in medication.

*Before the coronavirus, the gym became my life and went there 3, 4, 5 times a week and was my structure*…*it helped me cope with my isolation* … *with coronavirus, it removed my structure. Just gone. And I don’t function well like that* … *As a person living on my own, it’s the lack of external structure that’s the most destructive thing*. [P6]

Some people reported anxieties about lockdown easing and life returning to ‘normal’:

*It’s spiked my anxiety thinking that things are going to get back to normal and it’s a bit soon* … *People are kind of acting like it’s over and that just makes me incredibly nervous going outside*. [P26]

There was no clear pattern associated with diagnosis. Participants described the negative impact of living with specific conditions and how this intersected with associated symptoms:

*It’s kind of impacting on* … *hearing voices and stuff … I can’t escape them, can’t go out really just to kind of get like fresh air and clear my head, and I think kind of ruminating thoughts that I guess you’d describe as paranoid thoughts have been getting a bit worse since the lockdown*. [P20]

*Anxiety isn’t something that I normally have, I might have it fleetingly, but in lockdown, my anxiety levels have just been so high, like actually the physical sensations of feeling fear, just absolute terror and fear. I had it when I went in the shop, and I had to actually come out the shop because it was so, so bad*. [P30]

Some people described a positive impact on their mental health; lockdown provided a relief from external pressures, allowed enjoyment of a quieter world, while slowing down helped ‘contain’ mental health symptoms and provided more time for activities and self-care. Some participants had adapted and developed coping strategies. There was also optimism and hope, with some reporting that they had valued trying new activities or ways of connecting with people and hoped to maintain this:

*[I’ve] been doing gardening, colouring in, and exercise in last couple of weeks. And also, weekly catch-up with friends, have set it as every Wednesday evening so is good to have something to look forward to*. [P32]

### Relationships and (dis)connections

Participants experienced changes to relationship dynamics due to lockdown: changing caring responsibilities, parenting new babies and home-schooling children alone; and anxiety around bringing the virus home to at-risk family members. Lockdown tended to push families towards extremes; some moved in together while others reduced contact with relatives they did not live with. These impacts heightened existing mental health difficulties. For some, existing therapeutic relationships were lost, either because the lack of privacy at home meant therapy was discontinued, or through losing informal peer support from friendships formed within services:

*I’ve really missed … seeing the friends that you make through the [therapy] groups, other people that have got a mental health illness*. [P12]

Like the general population, participants connected in new ways during lockdown. Online communication seemed particularly useful for various faith and community groups:

*I’m part of a gay man’s community… and have created an online event for men to get together… we are talking about and planning on meeting in person in the future*. [P40]

Some people’s difficulties prevented online connection, especially video calls which two participants said triggered paranoid thoughts. Most interviewees reported increased loneliness that affected their mental health, even if living with others. Some felt *‘touch deprived’* [P40]; others noted that imposed isolation could be frightening or lonely in a way that chosen solitude was not. Some found it hard to discuss their problems during a universal crisis:

*I couldn’t talk to anyone about my personal problems in full because everybody was coming with their own problems. Everybody, my social worker was feeling the pressure and everyone else I knew*. [P48]

Some benefited from increased support from family members spending more time at home or from new connections made:

*There’s people coming up and down my road all the time. It’s really nice to see so many people … People have been a lot more neighbourly and, you know, communicating*. [P25]

However, while some formed new relationships through the weekly ritual of the communal ‘Clapping for Carers’ that took place from March to May 2020 in the UK, others found the ‘blitz’ spirit rhetoric hollow alongside long-standing social divisions:

*Seeing my neighbours every Thursday at 8 o’clock and giving a thumbs up to them … they know I’m alright and I know they’re all right*. [P11]

*There have always been elderly people, and people with mental and physical health difficulties struggling … why are we only taking it seriously now?* [P4]

### Unequal impact

Some participants explained how many of the impacts were exacerbated or felt unequally in relation to their race, culture, socioeconomic status or disability. Feelings of isolation and disconnection were intensified because participants were no longer able to connect with community. Some highlighted the impact of racism and stigma, noting that lockdown coincided with the Black Lives Matter protests of May 2020 and the increased media focus on systemic racism. Participants explained that increased risks in relation to COVID-19 for people from BAME communities intensified their fears and anxieties:

*With the news that people from Black and ethnic minority communities are more likely to get it, and die from it … a fear of a lot of people has been that we’ll go into hospital, and they’ll look at me and say ‘you’re not really worth that much, and we just won’t give you a ventilator if we don’t have enough’*. [P17]

Some BAME participants told us they had experienced increased racism and these experiences increased feelings of worthlessness and anxiety:

*People have been rude and nasty, they think they will catch it if they stand next to you* … *I was already in that margin of people who were stigmatised, and now we are all stigmatised*. [P17]

For one participant this had a profound effect on her mental health as it *‘triggered past memories of discrimination’* [P37] and reminded her of previous trauma.

Being in lockdown with family or close community posed a challenge where people felt their mental health problems were not understood. They felt judged and stigmatised by relatives and community, and without usual support from services this compounded feelings of disconnection and worsened mental health problems. The intersection of mental health difficulties with specific religious needs intensified some impacts. One Muslim participant moved in with family during lockdown, but was on a specific medication timetable so had to get up while fasting family members slept, exacerbating the tensions of close-quarters living.

The shift to remote services and online therapy was a barrier for several participants as their disabilities or finances limited access to technology. Lack of privacy further exacerbated tensions in families where mental illness was not accepted. A few participants noted that services were not culturally competent and did not account for the impact that race and culture would have on individuals during COVID-19, thus limiting access to appropriate support:

*They didn’t have cultural training, wasn’t their fault, but it felt alien to me* … *we live in a multi-cultural Britain* … *there should be more representation in services*. [P4]

The restrictions on activity during lockdown and the impact on health services exacerbated the mental health impacts of COVID-19 for those with co-occurring physical health issues, especially those with life-threatening conditions who had their treatment suspended:

*I have a physical health condition as well as mental health, but I still used to try and get out as much as I could* … *I find it really hard not to do the things that were part of my routine*. [P23]

*I was in a bloody panic* … *this is a very worrying time for those of us with cancer because all your scans are cancelled* … *hip operation and physio cancelled … [I’m] worried I’m making it worse*. [P6]

## Discussion

### Main findings

Our study identified five themes describing the experiences of people with pre-existing mental health difficulties during the COVID-19 pandemic. These captured difficulties in coping with day-to-day functioning, specific psychological impacts of the pandemic, struggles with social connectedness, and inadequate access to mental health services. Findings convey a sense that these factors had exacerbated existing mental health difficulties for many during lockdown. For a sub-set of individuals, aspects of their mental health improved in the absence of some of the stressors of pre-pandemic life.

People with pre-existing mental health conditions experienced serious disruption to their access to, and the quality of, mental health care as a result of the pandemic. The opportunities and challenges of remote mental healthcare were an important aspect of our findings. While for some people, telephone and digital support provided continuity of care, for others there were issues around access to technology, maintaining therapeutic relationship remotely, and digital interfaces exacerbating difficult feelings or symptoms associated with their mental health.

Inequalities in impact were widespread, including BAME groups’ fears about poorer health outcomes, not receiving treatment equitably, experiences of stigma resulting from perceived association between ethnicity and COVID-19, and direct experiences of racism and the trauma which that invoked.

Many individuals responded to difficulties in accessing statutory mental health care by developing their own coping strategies, reporting positive experiences in finding new sense of purpose, turning to other sources of support including the voluntary sector, and finding new ways of connecting to others and to community.

### Findings in the context of other studies

Our findings build on the international literature describing early impacts of the pandemic on people with pre-existing mental health conditions, detailing deteriorations in symptoms, experiences of loneliness and social isolation, and lack of access to services and resources, but also accounts of resilience, effective self-management and peer support, [10] also reflecting our participants’ concerns about the sustainability of alternative forms of support. Our findings complement research exploring the experiences of mental health staff during the pandemic, [16] also highlighting challenges and opportunities around new remote care. Our data hint at differential effects of COVID-19 for people with different mental health conditions, as reflected in the wider literature. [4,9,12]

### Strengths and limitations

Embedding lived experience into study design and conduct was a strength of our approach to understanding people’s experiences of mental health during the pandemic. Interviews conducted by researchers with lived experiences can enhance disclosure, [27] while the central role of lived experience in the analysis process, and particularly in understanding inequalities of impact, further enhanced the validity of our findings. [21]

Purposive sampling achieved demographic diversity, including people with a range of mental health conditions and experiences of mental health services. While we were proactive in exploring experiences of BAME people, we recognize this is not a homogenous group: the experience of different social and clinical groups requires more nuanced exploration in future studies. Because we recruited online our sample under-represents digitally excluded people whose experiences are particularly important to understand.

### Practice and policy implications

COVID-19 has reminded us that social determinants, including poor housing, continue to impact mental health and increase demand on services. [28] The pandemic also disrupted people’s self-care and access to essential physical health care. The role of mental health services in ensuring that people receive good physical healthcare has never been more important.[29]

Increased use of remote, and especially digital, mental healthcare is advocated by governments and healthcare professionals. [14,18] Lack of privacy in the home to engage in therapy or conversations with mental health professionals was prominent in our data. Consideration might be given to providing technology to enhance privacy, or prioritising individuals for face-to-face care where these issues are insurmountable. Particular attention might also need to be paid to forming new relationships remotely at the start of treatment or therapy. [16] Remote working in early response to the pandemic might not offer longer-term solutions; service providers should work with the people who use their services to offer flexibility in the range of available approaches to remote care. [18]

Mental health services might do more in providing, or connecting people to other services which provide, social interventions that specifically support building social connectivity [30] at this particularly challenging time. A lack of awareness of the cultural context to people’s experiences of the pandemic represents an additional systemic challenge in providing equitable mental healthcare to people from BAME communities that needs to be better understood and addressed in future service provision. [7]

### Future research

Our interviews were conducted between May and July 2020 when COVID-19 restrictions and the public’s responses were still evolving. Experiences and concerns may change, with further work needed to explore the cumulative impacts of continued uncertainty around COVID-19, the maintenance or tightening of social distancing measures, and ongoing challenges in the provision of mental health care. Follow-up studies, capturing both in-depth exploration of people’s experiences and validated outcome measures, are necessary, complemented by analyses of large routine datasets to understand the effects on specific groups over time. [18] Research is also needed to better understand digital exclusion for people for whom access to or use of technology is a substantial barrier, [31] and to understand how the digital therapeutic relationship can be built and sustained.

## Conclusions

Our co-produced, qualitative study identified the difficulties reported by people with pre-existing mental health problems during the COVID-19 pandemic in the UK, relating to coping with day-to-day functioning, psychological impacts of the pandemic, struggles with social disconnectedness, and inadequate access to mental health services. We identified experience of inequality of impact by those in marginalised groups defined by ethnicity, disability, and socio-economic status. Our findings accord with international literature; that the pandemic has exacerbated existing mental health difficulties for many during lockdown, whilst offering a sub-set of individuals relief from some of the stressors of pre-pandemic life. The experiences described reflect the relatively early days of the pandemic in the UK, but highlight the need for planned, sustainable, evidence-based adaptation of service provision to minimise further harmful impacts. We suggest that experiential knowledge and service co-design should be at the forefront of that endeavour, with service providers and researchers working in partnership with community organisations and people who use mental health services.

### Lived experience commentary by BC and TK

We participated in this study as Lived Experience researchers (LERs) and were satisfied by this partnership of true co-production which enhanced our qualitative research skills. We also had a weekly peer-led reflective space which proved a valuable forum for mutual learning and support. This involvement also boosted our self-confidence and general wellbeing. The time spent on the research benefited us during the pandemic as this was a valued and enjoyable activity.

Utilising LERs provided a deeper understanding of the interviewees’ experiences which helped guide the interview, and also provided a more natural rapport making the interview feel more conversational.

During interviews, we noted similarities in terms of our lockdown experience and the experience of participants, such as negative psychological impact due to lack of face to face contact, particularly social contact, but also clinical contact.

The study has revealed that there were people who seemed to be coping or even feeling better during lockdown. Given that therapy may disturb difficult issues, not having sessions or having them in a less intense way might be associated with feelings of wellness due to not being challenged. Similarly, with disorders such as social anxiety or agoraphobia, the easing of pressure translates to loss of resilience and hard-won progress. This could have long term negative effects.

Although this paper highlights some of the self-management strategies developed by most of the participants during the lockdown, we are questioning how sustainable these are without regular support, and longer studies would be merited. The paper also failed to fully capture the perspective of those who were digitally excluded or those excluded by disability or language barrier.

Ethnic minority groups reported significant anxiety around the virus due to disproportionate mortality. The compounded effect of such stress upon existing mental health challenges of diverse populations should be studied further.

## Data Availability

Interview transcripts are not publicly available to ensure individual participants are not identifiable in accordance with participants' consent and study ethical approvals. A table of additional illustrative quotes for each theme and the interview topic guide are provided as supplemental material.

## Ethics approval

Ethical approval for a study focusing on loneliness was obtained from the UCL Research Ethics Committee on 19/12/2019 (ref: 15249/001). An amended topic guide covering experiences of COVID-19 was approved on 04/05/2020.

## Consent to participate and to publication

All study participants consented to take part in the study and for anonymised quotations from their interviews to be used in publications.

## Availability of data and materials

Interview transcripts are not publicly available, to ensure individual participants are not identifiable, in accordance with participants’ consent and study ethical approvals. A table of additional illustrative quotes for each theme and the interview topic guide are provided as electronic supplementary material.

## Code availability

Not applicable

## Author contributions

All authors contributed to study design, data interpretation and writing the paper.

CD, JH, PN, RRO, PS, MB, UF, JO, EP and TS contributed to data collection.

SG, CD, JH, PN, RRO, PS, MB, UF, JO, EP, TS and BLE contributed to data analysis.

The NIHR Mental Health Policy Research Unit COVID Coproduction Research Group comprises the authors of this paper and: Katie Anderson, Nick Barber, Anjie Chhapia, Beverley Chipp, Tamar Jeynes, TK, Jo Lomani, Karen Machin and Kati Turner. KA, NB, AC, BC, TJ, TK, JL and KM contributed to study data collection and/or analysis. BC and TK wrote the Lived Experience Commentary which accompanies this paper. AC, TJ and KT, together with study author JH, facilitated regular meetings of a reflective space for study Lived Experience Researchers during the project.

