## Supplementary Material 1 for "Experiences of living with mental health problems during the COVID-19 pandemic in the UK: a coproduced, participatory qualitative interview study"

**Title of Study:** Exploring the lived experiences of loneliness and isolation with people with mental health problems during the COVID-19 pandemic in the UK.

**Revised Interview Topic Guide**

**<u>Introduction</u>**

Thank you so much for agreeing to be interviewed.

As you know the purpose of the interview is to find out about your experiences during the virus outbreak, and more generally, your experiences of feeling lonely or isolated and how they may relate to experiences of mental health problems. This will help us understand more about mental health, isolation and loneliness and how it relates to the virus outbreak. It will also help us to do further research looking at how to measure these things, or to develop ways of supporting people's mental health and reducing loneliness and isolation particularly during virus outbreaks. We will also learn more about the other challenges you have faced during the virus outbreak and what can be helpful for people experiencing mental health problems during this new situation.

There are no right or wrong answers, as everyone's experience will be different. What you say will be kept completely confidential and we'll anonymize the information you give us, so we won't use your name or say anything that could identify you, especially in anything we write. Please be as open as you feel comfortable, and if you want to skip a question or take a break at any point, please let me know.

Firstly, I'd like to ask you for some information about yourself, so we can understand the context of your answers in relation to the virus.

- Are you currently using mental health services **Y/N**
- If no, have you ever? **Y/N**; when was your most recent contact?
- What is the current/most recent types of service you have used?
- Who do you currently live with: **partner, children, other family, flatmates, alone**. (select all that apply)
- Has this changed due to COVID? **Y/N** (if Yes: who did you live with before)
- Are you in paid work? **i) Yes, currently working; (at home; or in a workplace?) ii) Yes but furloughed (currently not working but having your wages paid through the government scheme and/or your employer); iii) No**
- If yes: **i) Full time; or ii) part-time**
- Are you volunteering? **Yes/No** (If yes, please describe)

- Are you in education? **Yes/No** (If yes, please describe)
- What region of the UK do you live in? **[1. North East, 2. North West, 3. Yorkshire and Humber, 4. West Midlands, 5. East Midlands, 6. East of England, 7. London, 8. South East 9. South West]**
  - What kind of environment do you live in: **City over 100,000, smaller city or town, village, countryside/rural**
  - The public are being asked to stay at home as much as possible (that is, only leaving the house for food shopping, exercise or work if you can't work at home). How long have you been limiting your social contact with others in this way? **Haven't been, or duration in weeks**
  - Have you had any COVID 19 virus symptoms, or have you been diagnosed with the virus? **i) Yes, definitely; ii) Probably but unsure; iii) Not as far as I know**
- Are you shielding at home on advice from government, or currently self-isolating because of your symptoms, or someone else's symptoms? **i) Shielding to avoid getting the virus; ii) self-isolating because of my symptoms; iii) self-isolating because of others' symptoms**

**1. Can you tell me about the main impact of the virus outbreak on you?**
Prompts: day to day impact on routine, who you see or have any contact with, practical issues, access to services and support, going outside and whether any difficulties arise when you do. Impact on relationships with family/friends.

**2. (For those using mental health services) How has the care you receive been affected by the virus outbreak?**
Prompts: what service did they receive before, i.e. how often did they see someone from service, was it face to face, was it in outpatients, or at home? What has changed, or stopped? Has it changed mid treatment/therapy? How have they found adapting to any

changes from face to face contact? Was there any choice or consultation about how the changes to your mental health care were arranged?

**3. Do you feel isolated or lonely currently, during this "stay at home", lockdown time?**
Prompts: Have you felt more or less lonely during the virus outbreak?
How does loneliness impact on you day to day during the virus outbreak/ and before the virus outbreak?

**4. Can you tell me about how your mental health has been since the virus outbreak has developed? What about since the lockdown "stay at home" advice has been put in place?**
Prompts: tell me more about the changes. Any things which have got more difficult, new difficulties? Any positive effects of the virus outbreak and staying at home on your mental health?

**5. How are you coping with your mental health in the current situation?**
Prompts: has anything helped? Has anything you tried, or any support you were offered, not worked? Any barriers to accessing new options available, i.e. when people live with lots of people, is privacy for phone calls to services difficult, finance to make calls.
What support or guidance on mental health and the virus have you read or heard about? (e.g. via social media or websites, from leaflets, or on TV, from any mental health charities like MIND, or NHS? What guidance have you followed? Have you found it clear, has it been helpful? [Explore: managing social distancing and knowing when you can go out; hygiene measures e.g. handwashing, wearing masks]

**Now I would like to ask you about experiences of loneliness in your life more generally, not just in relation to the current virus outbreak.**

**<u>6. Experience of loneliness</u>**
**Can you tell me what the word lonely means to you? How would you define it?**
Prompts: How does it make you feel? Physically, psychologically, emotionally e.g. social anxiety.

**7. Has there been a time or times in your life when you have been lonely?**
**Would you say you feel lonely in your life in general at the moment?**
**What is/was that like?**

Prompts: Are there things that you think have triggered or underlie your feeling of loneliness? E.g. age, culture, personality style, , difficulty fitting in, stopping work, moving location, loss of a partner or family.
Are there situations or times when you feel more lonely than others? Prompt: e.g. seasonal, not working.

**Social contact and loneliness**

**8. Do you feel lonely when you are in the company of others?**
**Are there any kinds of social contact that make you feel more or less lonely**?
Prompts: E.g. groups, family, special friend, in person rather than online or using technology. When you meet people in person, do you feel that you can talk to them easily?

**9. Do you think that spending time on your own can be helpful/therapeutic?**

**10. Does spending too much time on your own negatively affects your wellbeing?**

- [If yes] - How much is too much? Where is the line?

**Loneliness and mental health**

**11. Do you think feeling lonely is connected to your mental health?**

- [if yes]: In what ways?

Prompts: Does feeling lonely make your mental health worse? In what ways? Do you think your mental health problems or your treatment contribute to your loneliness? [Prompts: e.g. side effects of medication, the age you developed mental health problems.]

- Thinking back to when your mental health problems first started, do you think that feeling lonely came before or after?
- [If loneliness came first]: Has your experience of loneliness changed since having mental health problems?

**12. Do you feel you belong in the community around you?**
Prompts: explore different communities: neighbourhood, family and friendship groups, communities of interest [If not]: Can you tell me more about this feeling of not belonging? In what ways, if any, does that relate to your feeling of loneliness?

**Final questions:**

**13. Are there ways in which you have tried to reduce your loneliness? What has and has not worked?**
Prompts: Do you have ways of coping? Have you had any support? What kind of support? Has this helped?

**14. What would not being lonely look like for you?**
Prompts: How do you imagine your life would be different?

**15. What tips or advice would you give someone who was struggling with loneliness?**

Finally, I’m going to ask you for some information about yourself. We are collecting this so that we can make sure we interview a range of people, and to help us provide some context to what you say. If there are any questions you would prefer not to answer, please just ask the researcher to move on to the next question: you don’t have to provide any information you don’t feel comfortable sharing.

- How would you prefer to describe your gender:

1. Male
2. Female
3. Another way (If in another way, please tell us what term you prefer)
4. Prefer not to say

- Do you consider yourself to be a trans person? **(Yes, No, Prefer not to say)**

- Age (in years)

- **How would you describe your ethnicity?**

**White**

- English / Welsh / Scottish / Northern Irish / British
- Irish
- Gypsy or Irish Traveller
- Any other White background

**Mixed / Multiple ethnic groups**

- White and Black Caribbean
- White and Black African
- White and Asian
- Any other Mixed / Multiple ethnic background

**Asian / Asian British**

- Indian
- Pakistani
- Bangladeshi
- Chinese
- Any other Asian background

**Black / African / Caribbean / Black British**

- African
- Caribbean
- Any other Black / African / Caribbean background

**Other ethnic group**
- Arab
- Any other ethnic group

- What is your first language?

- Which of the following best describes your sexual orientation:

- **i) Heterosexual/straight; ii) Bi/bisexual; iii) Gay/lesbian; iv) Prefer not to say**

- Do you consider that you have a disability **Y/N**

- Do you take medication prescribed for mental health condition(s) **Y/N.**

Version 2: 16/04/2020
