## Supplementary Material 2 for "Experiences of living with mental health problems during the COVID-19 pandemic in the UK: a coproduced, participatory qualitative interview study"

**Supplemental Table with details of subthemes and additional quotes**

**Steven Gillard[1], Ceri Dare[2], Jackie Hardy[2], Patrick Nyikavaranda[2], Rachel Rowan Olive[2], Prisha Shah[2], Mary Birken[2], Una Foye[3], Josephine Ocloo[3], Ellie Pearce[2], Theodora Stefanidou[2], Alexandra Pitman[2], Alan Simpson[3], Sonia Johnson[2], Brynmor Lloyd-Evans[2]**

**On behalf of the NIHR Mental Health Policy Research Unit COVID Coproduction Research Group**

The NIHR Mental Health Policy Research Unit COVID Coproduction Research Group comprises the authors of this paper and: Katie Anderson, Nick Barber, Anjie Chhapia, Beverley Chipp, Tamar Jeynes, TK, Jo Lomani, Karen Machin and Kati Turner.

1. Centre for Mental Health Research, City, University of London, 1 Myddelton Street, London EC1R 1UW

2. Division of Psychiatry, University College London, Maple House, 149 Tottenham Court Road, London W1T 7NF

3. Institute of Psychiatry, Psychology and Neuroscience, King’s College London, 18 DeCrespigny Park, London SE5 8AF

**Supplemental Table with details of subthemes and additional quotes**

| Theme | Subtheme | Brief Description | Illustrative quotations |
|---|---|---|---|
| **Impact of COVID-19 on everyday life and mental health** | **Homelife** | Difficulties with access to shops and essential goods and services | *''Because I do not have a car it has been an absolute nightmare to get food - my friend has been helping me by bringing me the food shop.''* **P35**<br>*'Businesses are not opening now. There are things I cannot get online'* **P38** |
| | | Disruption to usual activities | *''I went out Monday evening and the beach was packed, there were like hundreds of people on the beach and swimming in the sea.. so mixed feelings. It often is the best part of my day but a part of the day that causes me stress, like anger as well''* **P3**<br>*'Lockdown meant you are very limited to do things you'll normally be doing'* **P38** |
| | | Overcrowding at home and lack of privacy | *''Privacy was an issue because for the* [peer support] *group, you cannot have anyone around, no one should be able to hear the group or see the group and attendees, so it meant I had to go about doing things differently"* **P29**<br>*"I'm quite lucky… I have a house and it's just me and my partner so I can get that privacy which I'm very grateful for."* **P33** |
| | **Employment** | Ending or disruption to employment or education | *'' I was only away from home for six months so I feel like I never got to experience [university] properly so that was pretty disappointing''* **P34** |
| | | Lack of access to requirements for work | *"I have issues I don't have all my equipment so it is more painful working from home"* **P13** |
| | **Finances** | Big impact of unemployment or lack of work activities on finances | *''I have been extremely short of money lately and I have just been dodging and diving''* **P13** |
| **Impact of Changes to Mental Health care** | **Changes to Mental Health care** | **How changes to mental health services and care have impact directly on** | *"Mind more therapeutic than NHS….more holistic so better recovery outcomes."* **P7** |

| Theme | Subtheme | Brief Description | Illustrative quotations |
|---|---|---|---|
| | | **the individual's well-being and mental health (positives and negatives)** | *"I usually get the same GP but can't at the moment…it's difficult opening up to new GPs if I need something."* **P5**<br>*"The Recovery College is not therapy but has been good to have the support"* **P21**<br>*"I was seeing her every week and it's been cut to every 3 weeks." (and to phone appointment)* **P25**<br>*"It [phone therapy] was okay to be honest, I felt possibly a bit more comfortable being at home."* **P26** |
| | **Remote Mental Health Care** | **Transition to digital/ remote care, including the barriers (e.g. privacy) and facilitators for individuals to manage this change to remote care (positives and negatives)** | **"***I was looking on line for some sort of alternative recovery methods. I like thing like yoga, meditation, art, music and poetry, so I was looking for these kind of supplements to the medication I am taking.* **P8**<br>*'Being able to talk about things is a lot harder when you're talking from home. In case people listen in or the call's not secure"* **P17**<br>~~*''It hadn't really occurred to me to look for something formal , like online support or anything''* **P2**~~<br>*'It was strange because it was like [psychiatrist] was right in front of me...but because you're at home, I feel I never invited you here Mr. Psychiatrist to my home!'* **P27**<br>*"I'm still in contact with them as much as I was before, it's just been predominantly phone based."* **P20**<br>*"Zoom meetings make people feel uncomfortable ... you start worrying about how you look."* **P6** |
| | **Prioritisation of services** | **Resentment that staff are being prioritised for mental health care ahead of those waiting for years** | *It's like they've done a certain job and are up here….it's like if you have a certain job title or qualification, you're recognised* **P4** |
| | **The changing relationship with non-NHS mental health support in** | **Complementary or alternative therapies; Voluntary or community / grassroots organisations bridging** | *"I was looking for these kind of supplements (complementary or alternative therapy) to the medication I am taking."* **P8**<br>*'The manager of my day centre has started doing home visits – he can't come into your home but he can meet you outside and take you for a walk – that was good.''* **P41** |

| Theme | Subtheme | Brief Description | Illustrative quotations |
|---|---|---|---|
| | **the community and beyond** | **the gap; people tried these out for the first time, or came to rely on them more, because they were limited in their access to formal or informal support** | *"I call Mind's crisis line every day and have made relationships with some of the call handlers"* **P43** |
| | **Medication** | **Information capturing how the pandemic has impacted on people's access or use of medication** | **"***Had medication increased-too much right now, especially as on more medication."* **P14**<br>*"They were very good, they got the prescription sent straight to the pharmacy so I could collect the medicine without having to be seen*." **P18**<br>**"***GP won't do shared agreement (with CMHT) [for change in medication prescribing] until I've been for an ECG, but I don't really want to go to hospital.* **P23** |
| | **Communication between services and the individual** | **Increased communication and willingness of healthcare staff**<br><br>**Lack of communication and confusion/ uncertainty** | *"Puzzle pack sent out by Trust...puzzles, activities to do." (*to service users who go to Recovery College) **P23**<br>*I never heard anything from them – I have had like no contact whatsoever... for a couple of weeks* **P36**<br>*'They did send a letter saying there was pressure on the service, and they may not be able to help as much but when you actually call them it is possible to speak to your care coordinator''* **P35**<br>'*Ward round was no longer ward round – consultant was going to people's rooms, which made me scared whether these new people had the virus'* **P42** |
| **Psychological impact** | **Direct impact on the individual** | Negative and positive experiences of covid and lockdown on existing mental health conditions and psychological well-being | *"Just my mood's been flat, and I'm a voice hearer, my voices have been worse, that's been quite difficult….I self harm and that's got worse."* **P12**<br>*''One of the problems is I would get very low during the day…. I haven't been in that situation during the last 6-8 weeks*" **P13**<br>*"I've actually really enjoyed it. I have actually found myself less isolated during lockdown than when everything is back to normal..this is the first time in about 10 years that I have has such good mental health...I have been able* |

| Theme | Subtheme | Brief Description | Illustrative quotations |
|---|---|---|---|
| | | | *to do quite a lot of reflecting, and have spent more time on my wellbeing than I normally do...“I am dreading when we come out of lockdown again”* **P25**<br>*“I was managing with my medication quite well but since Covid arrived that has sort of changed and I have had more issues with my mental health because of it.”* **P35**<br>*“Slowing down has forced me to confront my demons...and open up more.”* **P43** |
| | **Resilience, self-care and personal strategies** | Use of various practical and psychological strategies to manage mental health, both active and avoidance of ‘negative’ factors.<br><br>Decline in personal self-care. | *“I haven’t been able to do activity planning like seeing people regularly I haven’t been able to do my usual coping skills that I was taught’’* **P34**<br>*“I’ve stayed away from social media , reading people’s posts...people’s negative thoughts...I watch the news once a day and only reads the government advice website so it’s not overwhelming.”* **P32**<br><br>*“Like not washing as much as I would if I was going out, not doing makeup, not doing my hair… because I live by myself and it’s just, like, mental effort”.* **P20**<br>*“It’s very hard for me to leave the house because I feel very tired and lethargic and like my self-care has dropped, I feel awkward to go outside because I haven’t showered.”* **P41** |
| | **Experience of stigma** | Experience of stigma and feeling judged relating to mental health, having to shield, financial issues, and being a single father. | *“From South Asian community, mental health misunderstood and stigma…family and community opinions...They see it as a way of life, not illness.”* **P7**<br>*“felt left alone with my son and establish a bond with him, for him to know I could do it*.” **P24**<br>*“I talk to my cousin a lot and he knows that I’ve struggled mental health wise but he’s self employed so he’s had a really hard time so I haven’t really felt comfortable talking to him about [my mental health]… it feels a bit selfish because he’s also not in a good place.”* **P26** |
| | **Changes in coping over time** | How people’s experiences changed | *“It’s got worse as things have gone on. In the beginning, not knowing how long it will go on for, so I was up and out….but now I feel less motivated*.” **P23** |

| Theme | Subtheme | Brief Description | Illustrative quotations |
|---|---|---|---|
| | | during the course of lockdown | *"As time's gone on it's definitely made me feel more anxious and a bit depressed being stuck indoors."* **P26**<br>*"Mental health has been very up and down. I've had a lot more going from manic happiness to depression and very few times of middling mood...has got more pronounced as lockdown gone on, though don't know if that's also die to other world events happening….the Black Lives Matter and most recently J.K Rowling being a bitch to trans people, and the GRA consultation, ignoring 70% people because of trans 'activists'...I am bloody furious and angry and it's really affected my mood"* **P27**<br>*"I think I've quashed some of my anxiety but, whether I have in reality, but I just feel a bit calmed myself maybe This is a coping strategy."* **P30**<br>*"[No change] I just continue to live my own life – keep to myself."* **P39** |
| | **Confusion and clarity** | Negative experiences of confusing guidance | *''It can be extremely anxiety inducing and provoking to read like pretty much anything about the outbreak and on the one hand you really wanna fell like you are on top of things and you are doing the right thing, and you are not accidentally flouting the guidelines because they changed two weeks ago and you forgot to check''* **P3**<br>*"My wife has a physical condition so it was unclear whether we both needed to be shielding"* **P9**<br>*"The government guidance was clear until they went and changed it. Stay alert is pretty meaningless for a virus. You can't see it coming.<br>and there's the specific anxiety, this idiot government, easing lockdown probably too soon and doing all sorts of other stupid things...I've been as annoyed as anyone else about the government's actions about Dominic Cummings and all that stuff, I've been following that."* **P11**<br>*"As things eased it's been uncomfortable… it's spiked my anxiety… I've been comfortable knowing what the rules are when they've been clear."* **P26**<br>*''Any time he was unwell, and we needed to call emergency services, they never came with any PPE - so it was like nothing had really changed."* **P36** |
| | **Fears about the virus** | Fears directly related to the virus and the | *"Because you are spending so much time on your own, you are thinking have I got it (the virus)? Has anyone else in the house got it? All these negative* |

| Theme | Subtheme | Brief Description | Illustrative quotations |
|---|---|---|---|
| | | psychological impact | *thoughts"* **P10**<br>*'The virus is still there and it is not just, you die and you could make someone die around you. It is a complete madness on top of my own, isn't it?'*. **P48**<br>*"When I go outside, [the voices] like say 'that person's going to infect you' or like say someone coughs they'll be like 'you're gonna die'."* **P20** |
| | **Hopes and uncertainty of the new normal** | People described positive experiences on their mental health and daily living as a result of covid-19 as well as hopes and anxieties about the new normal | *"We had a video call for first time...has opened up new things I hadn't tried...More aware of self-care strategies and things that are good for me. Feels the "Freedom to be myself."* **P4**<br>*''It's been a very good way of getting involved with people so I do not have to feel so lonely''* **P13**<br>*''I have picked up exercise for the longest time in my adult life now… been helpful to my mental health..I have been reading a lot more, which I think is good for your mental health...It has made me more serious about managing my mental health"* **P29**<br>*'...I have been self-isolating for a while and things change and I change for the better'* **P38**<br>*"We create our own happiness...there's a lot you can do to combat loneliness…I'm optimistic about the future."* **P43** |
| | **Anxieties and Fears related to changes brought on by Covid-19 and (end of) lock down** | A range of emotions were described by people that were directly a result of the changes brought on by Covid-19, the lockdown, and the relaxing of some rules during lockdown. These ranged from anxiety, fear, and guilt and were prompted by actual changes or anticipated changes | *"I watched the Horizon documentary about the pandemic, Tuesday of last week I think it was, and that said it could go on forever and I thought… there's an anxiety it will go on for a long time"* **P11**<br>*"when we do socialise again that's going to be trickier for me than it would've been, although I feel comfortable being at home kind of hiding from the world...I felt I was the only one keeping my distance… it just made me feel nervous about, well, entering the job market again for example."* **P26**<br>*"we don't know how it's transmitted or who is at risk. But the guidance put 'the fear of god in you' about it being on letters and parcels."* **P30**<br>*"I feel guilty that I haven't been able to see my grandma and be more supportive to her because she lives alone. But at the moment that is not happening and it is not on the cards for the foreseeable future so I feel bad"* **P35** |

| Theme | Subtheme | Brief Description | Illustrative quotations |
|---|---|---|---|
| | | | *“I felt that mental the health services they did really failed people with, you know, the clients really, because for me what could kill me first could be my mental health before any pandemic or anything...Sometimes I was in a crisis and I just didn’t want to call anyone in case they take me to hospital and I was scared for me to end up in hospital, because of the pandemic; everything was kind of a nightmare.”* **P48** |
| | **Purpose** | People described both positive and negative experiences of having routine and structure during lockdown as well as the negative impact of losing pre-lockdown activities and structure, including, employment, volunteering, and leisure and social activities. For many people, this led to a decline in mental health. During lockdown, people spoke about developing new routines and finding new purpose, such as childcare and cooking. | *‘’I think it definitely has helped being conscious of what I eat, planning, like going to the shop is obviously the highlight of my week, it’s like almost a social event (but not really), planning what I’m eating, cooking nice food, baking, trying to keep my flat looking nice like buying flowers and stuff’’* **P3**<br>*“Everyday would go to cafe in morning for coffee and read newspaper, and then do food shopping. Go home , then back out to big shopping centre. Go home then out again to cafe. Home for dinner and then another walk for 8 to 9 miles a day before lockdown...haven’t bought a newspaper or had a coffee or been o shopping centre. Don’t miss it, but my day has changed.”* **P4**<br>*‘’I have tried to do my best to help where I can I have been pushing the issue of BAME communication’’* **P9**<br>*“Not having as much social support or distraction has kind of intensified* [voice hearing] **P20**<br>*“Doing things around house and normal routine was difficult because there was no structure to day or week….just killing time. Not aware of what day it was and fell apart...Since being back at work for 3 weeks, this has helped a lot.”* **P22**<br>*“Purpose now is caring for young son and working on developing a bond with him...felt left alone with my son and establish a bond, for him to know I could do it...frustration having to do it alone.”* **P24** |

| Theme | Subtheme | Brief Description | Illustrative quotations |
| --- | --- | --- | --- |
| | **Vulnerability of isolation** | Different ways in which people experienced isolation and loneliness. This included people living alone and those who had people around them. People described how remote contact was no substitute, nor having to do usually social activities alone. Some people talked about how being isolated at home reminded them of other periods of social isolation due to mental health reasons. | *"You thinking you are on your own and you have nobody else to express these negative thoughts to. So, you keeps all those negative thoughts to yourself and of course it spirals.''* **P10**<br>"*felt like a prisoner in home, as cannot see people..because of shielding cannot go out and talk to neighbours*" **P14**<br>"*I was struggling to socialise before it happened and.. going out and exposing myself to social situations was the healthy thing to do … the lack of that has made me feel more nervous going out*." **P26**<br>*"I think it gets me a bit low because I'm very much aware that I am lonely, it's just literally always there- it's just like it's becoming a pandemic loneliness"* **P29**<br>"*I can't stop thinking of things a little bit more, I was diagnosed (with a mental health condition) a couple years ago and I didn't leave the house for six months and during lockdown and feeling isolated has reminded me of that time when my mental health was at its worst...prior to lockdown I hadn't been thinking much about it but now I'm thinking about it a lot more and it's just been backlog memories from that time''* **P34**<br>*"I can't go to the gym at the moment because they are closed as such, so, I do some of the exercises at home, but it is difficult, but it is difficult. I find difficult to do things on my own because I don't find the same level of enjoyment as when it is a social activity*" **P49** |
| | **Impact of physical health concerns** | Psychological and practical impact of physical health conditions on living through lockdown and feeling particularly isolated, especially for those that were shielding. Feelings of anxiety and frustration from not being able to | "*More recently I have hit a low because I didn't want to go out and I put it down to Covid and lockdown and I think that's been the biggest impact for me because it just made it really hard…to look at what's going on and take those steps if I need to earlier rather than working till I'm burnt out and then having to take a longer period of sick leave which I haven't had to do for quite a while but unfortunately I need to do now because of my mental health."* **P35**<br>"*Hard on my physical health as there are no appointments. Impacts me going out and doing activities. If you don't feel well physically, affects mental health. Feel like I'm dealing with everything myself*." **P4**<br>*"I just can't deal with the isolation [of shielding], living by yourself isn't the* |

| Theme | Subtheme | Brief Description | Illustrative quotations |
|---|---|---|---|
| | | access usual physical healthcare, especially in those with serious health conditions, and anxiety and fear around catching covid whilst living with an 'at-risk' condition. | *best during a lockdown."* **P20** *"I was in a bloody panic...this is a very worrying time for those of us with cancer because all your scans are cancelled...hip operation and physio cancelled. Have no knowledge [of how to do the exercises] and worried I'm making it worse."* **P6** *" have thyroid problems, I have got hypothyroidism, so, sometimes I don't have energy at all, Sometimes I get very low in my mood. So, I couldn't be active or be, you know, thinking in terms of how making contribution to different things"* **P48** |
| | **Impact on recovery** | Impact of lockdown on usual use of positive coping strategies to assist recovery | *"I have PTSD… not having a good time with that in February and that then triggered the depressive episode. And usually when I'm recovering from that sort of thing, I try and socialise more and that brings me out of it."* **P5** *"I think there would have been less anxiety and more ability to help myself recover if lockdown hadn't restricted what I usually have when I recover from that sort of episode."* **P5** |
| **Relationships and (dis)connections** | **Pros & cons of being around family more** | Increased closeness / time spent indoors with family brought both benefits and challenges to participants: some benefitted from having someone to count on, while others struggled with lack of privacy. | *"Living with parents helped me, they do weekly shop, cook meals and help me manage."* **P7** *"I try and ensure the doors are shut so that I do have a bit of privacy [to speak to care coordinator or psychologist-] but it's just impossible sometimes, when someone just comes walking into the lounge… it makes it very hard"* **P12** *"I am the kind of person that needs to be alone occasionally and be independent and do my own thing. Feeling like there is always somebody there is quite difficult for me in terms of anxiety and some of my other issues."* **P19** *"I'm quite lucky because my partner is really supportive and does help me while I'm at my worst."* **P35** |
| | **Caring for others** | People with mental health problems were both carers and cared-for during lockdown. | *"I was supporting my friend as well and it was more distressing and I worry about people I love and care about. I wasn't worried about my personal safety, because I am already used to my symptoms, my stress. So, I have been trying to keep strong for everyone else, just wishing for the best for the people I love and care not to become ill."* **P46** |

| Theme | Subtheme | Brief Description | Illustrative quotations |
|---|---|---|---|
| | **Parenting pressures and positives** | Lockdown and the pandemic brought some unexpected positives in parent-child relationships, but also a huge amount of stress with managing appropriate information for children to access. | [My daughter] "*was obsessed, when they kept putting the figures out about 'today the youngest person who's died was 12 and they had no underlying conditions', she became obsessed with, you know, the youngest person who'd died that day without an underlying condition and that's the hard evidence that we were presented with through the media and as a parent, you can't separate emotion from that fact."* **P30**<br>*"The good thing has been that I have been able to support my daughter… So, that has been something I've enjoyed and to be involved in her education I suppose, to be able to be supportive*. " **P49** |
| | **Family disconnection** | Not having family physically around, e.g. family abroad or estranged, not being able to rely on grandparents for childcare. | "*Having not seen her for a prolonged period, It is starting to get to me now (not seeing mother physically)"* **P35**<br>*"I have been away from my family and they were a protective factor, buffer, safe space."* **P29** |
| | **Community spirit & connections** | Impact of the pandemic on sense of community: local, religious, etc. Impacts could be positive or negative, with increased connection but also increased judgement from neighbours and frustration from some participants. | "*The whole 'Stay happy' message and likening it to World War 2... Feels false."* **P6**<br>*"The Salvation Army have been bringing food parcels to my door, I have been in touch with them and they have been in touch with me so that's been very good. Same thing with Age UK… they have been turning up with amazing food parcels and I just feel very blessed"* **P13**<br>"*At one point I was talking to [a friend] from my doorstep as she was stood next to her car, we were having a catch-up, it was well over two metres away, we just felt judged so she wanted to go cos [the neighbours] were all staring at her."* **P35** |
| | **Isolation** | Physical and emotional isolation increased due to the pandemic & lockdown. | "*Since March, I have cried every day and I'm not normally emotional. It's a year's worth of sadness. There's a loneliness to having to sit with distress by myself… A nurse gave me a hug when I was in hospital and now I can't have that."* **P4**<br>*"Even living with other people in the house and pets, I still felt quite alone."* **P9** |

| Theme | Subtheme | Brief Description | Illustrative quotations |
|---|---|---|---|
| | | | *"I didn't necessarily go out much before the lockdown but I think the fact that you can't see people… has a toll on you."* **P20**<br>"*Main impact is not being able to hug people, being touch deprived...I'm normally a tactile person so is the worst thing for me."* **P40** |
| | **New ways of connecting** | Increases in virtual or online connections and the pros and cons of these, as well as challenges they brought such as digital exclusion and screen fatigue. | *"I have a weekly contact to read stories to my grandchildren on Monday evenings by zoom."* **P11**<br>"*A friend and I chat on Discord whilst watching an anime series together."* **P24**<br>*"My church small group has helped – we have a WhatsApp group and we speak every Wednesday.* "**P35**<br>*"My family tried doing WhatsApp or Skype or something and like "Oh let's do a video chat with the cameras on!" and I had to unsubscribe because I found it too difficult to manage participating because of long standing issues with video screens and that kind of thing… related to my more schizo type symptoms*." **P44**<br>"*For my paranoia, it [video calls] makes it worse, so I tend not to do them*."**P17** |
| **Unequal Impact** | **Increased risk and fear for specific groups** | Experiences of increased fear and risk, and feelings of stigma from others due to being in a higher risk covid-19 group, particularly people from BAME backgrounds | "*With the news that people from Black, and Ethnic minority communities are more likely to get it, and die from it, and the DNRs that have been popping up on people's homes, I've been waiting for mine. A fear of a lot of people has been that we'll go into hospital, and they'll look at me and say, you're not really worth that much, and we just won't give you a ventilator if we don't have enough."* **P17**<br>'*This virus is more deadly to us than a lot of other people*.' **P17**<br>"*Lots of my friends have lost a lot of people - The whole BAME community, the whole thing is awful – we've lost a lot of staff at work but people are not talking about it*" **P11** |
| | **Racism/ Black Lives Matter Specific** | Experience of attending or wanting to attend recent Black Lives Matter protests and impact of racism during the pandemic | *"I just wanted to be more involved, but, you know, I couldn't go to demonstrations or anything. And at the same time, my energy level was so low, I have been quite unwell for some time…… and I couldn't do that basically, my energy levels and I was too worried about everything*" **P48**<br>"*People are putting their lives at risk during Covid for people's fundamental freedom and for people to not understand that, that's been really painful*" **P36** |

| Theme | Subtheme | Brief Description | Illustrative quotations |
|---|---|---|---|
| | | | *''I went to one of the protests in my area and like it was really good and that helped me because there was a lot of fear around like who - the people that are silent, maybe they hate us, hate me as well but I saw those same people I was worried about being silent and I saw them so I was like there isn't, so it's not that bad and that definitely helped my mental health''* **P36** |
| | **Cultural challenges and supports** | Theme exploring the additional challenges and supports that culture has on individuals during covid-19, and the impact that these cultural aspects have on individual's mental health.<br><br>Examples of these include:<br>the impact of changes in local mental health care provision, e.g. the new service not taking into account of the cultural background of population; translation service; etc;<br>the impact that living in lockdown with family where culture does not understand mental illness has on the individuals | *"English is not my first language...so when I'm looking at groups...I need a group that spoke my language that I could sit in and be able to understand what was going on with my peers...Instead, I have groups where people are not from a similar background and they don't understand what I'm trying to say….It makes you feel further isolated."* **P17**<br>*"I'm a practising Muslim and when we have a funeral, we have certain rituals, so now we weren't able to do that [when family member passed]….sense of community gone."* **P17**<br>*''We don't really discuss mental health at all even though I would say there are other people in my family who have had mental health issues, it's just not talked about, so when I start having these issues and need someone to talk to , I still don't in my family''* **P34**<br>*"Very few people speak English here and there wasn't any appropriate information on coronavirus...so a lot of people just mistook it as a type of 'flu, didn't know about transmission"* **P17**<br>*"From South Asian community, mental health misunderstood and stigma … family and community opinions … They see it as a way of life, not illness."* **P7**<br>*'The doctors are Asian [in his area] but they are more westernised...my thinking comes from a more Easternised point of view and they don't understand that.'* **P17** |

| Theme | Subtheme | Brief Description | Illustrative quotations |
|---|---|---|---|
| | | the impact of being disconnected from cultural community | |
| | **Remote technology use** | Theme exploring the unequal impact that moving to remote technologies for services has on those from lower SES, those with disabilities and those with cultural differences not acknowledged during these moves. | "*Peer online groups are hard as many people are technophobic*." **P17** |
